# SARS-CoV-2 Seroprevalence Rates of Children in Louisiana During the State Stay at Home Order

**DOI:** 10.1101/2020.07.07.20147884

**Authors:** Monika L. Dietrich, Elizabeth B. Norton, Debra Elliott, Ashley R. Smira, Julie A. Rouelle, Nell G. Bond, Kéren Aime-Marcelin, Alisha Prystowsky, Rebecca Kemnitz, Arunava Sarma, Sarah Talia Himmelfarb, Neha Sharma, Addison E. Stone, Randall Craver, Alyssa R. Lindrose, Leslie A. Smitley, Robert B. Uddo, Leann Myers, Stacy S. Drury, John S. Schieffelin, James E. Robinson, Kevin J. Zwezdaryk

## Abstract

**BACKGROUND:** Children (≤18 years) account for ∼20% of the US population but currently represent <2% of coronavirus disease 2019 (COVID-19) cases. Because infected children often have few or no symptoms and may not be tested, the extent of infection in children is poorly understood.

**METHODS:** During the March 18^th^-May 15^th^ 2020 Louisiana Stay At Home Order, 1690 blood samples from 812 individuals from a Children’s Hospital were tested for antibodies to severe acute respiratory syndrome coronavirus 2 (SARS-CoV-2) spike protein. Demographics, COVID-19 testing, and clinical presentation abstracted from medical records were compared with local COVID-19 cases.

**RESULTS:** In total, 62 subjects (7.6%) were found to be seropositive. The median age was 11 years with 50.4% female. The presenting complaint of seropositive patients was chronic illness (43.5%). Only 18.2% had a previous positive COVID-19 PCR or antibody test. Seropositivity was significantly associated with parish (counties), race, and residence in a low-income area. Importantly, seropositivity was linearly correlated with cumulative COVID-19 case number for all ages by parish.

**CONCLUSION:** In a large retrospective study, the seropositivity prevalence for SARS-CoV-2 in children in Louisiana during the mandated Stay At Home Order was 7.6%. Residence location, race, and lower socioeconomic factors were linked to more frequent seropositivity in children and correlated to regional COVID-19 case rates. Thus, a significant number of children in Louisiana had SARS-CoV-2 infections that went undetected and unreported and may have contributed to virus transmission.

## Introduction

In December 2019, novel pneumonia cases were reported in Wuhan, China.^1^ On March 11, 2020 the World Health Organization (WHO) declared severe acute respiratory syndrome coronavirus 2 (SARS-CoV-2) a global pandemic. As of July 6th, 2020, there are over 11.6 million confirmed cases accompanied by over 537,000 deaths worldwide. Clinical manifestations of the infection range from severe viral pneumonia with acute respiratory distress syndrome (ARDS), to asymptomatic, with estimates of asymptomatic individuals potentially as high as 20%.^2^ Most children diagnosed with coronavirus disease 2019 (COVID-19) have been reported to recover in 1-2 weeks and have milder symptoms, faster recovery, and a better prognosis compared to adults.^3,4^ Among US cases reported between February 12th and April 2nd, 2020, only 1.7% of cases were in children aged <18 years.^5^ While healthy children do acquire SARS-CoV-2 and can become severely ill, this is less likely to occur than in adults.^6,7^ To date, it is unclear whether children are less readily infected, or whether they simply hold a higher probability of experiencing asymptomatic or mildly symptomatic infection and are therefore not tested.

SARS-CoV-2 shed by asymptomatic, pre-symptomatic, or mildly symptomatic subjects has likely contributed to the rapid spread of the virus.^8^ This is supported by contact tracing to pre-symptomatic or asymptomatic individuals, evidence of high viral shedding in these individuals^9^, and epidemiologic models.^10^ Symptomatic and asymptomatic children may act as efficient vectors of transmission, particularly within households, as high level shedding has been demonstrated in infected asymptomatic children.^11-13^ Currently, little is known about how children may contribute to broader spreading outside of households, a critical factor in guiding future infection control measures and informing predictive models of viral spread.^14^

Given the expected prolonged course of the pandemic, and the on-going efforts to balance recommendations for social distancing with the educational and developmental needs of children, there is a critical and urgent need to understand the prevalence of infection in children, and their roles in viral transmission. Here we describe evidence of previous infection in children in the New Orleans area that likely preceded the first documented cases of COVID-19 in pediatric patients.

## Methods

### STUDY OVERSIGHT

The study was performed in accordance with Good Clinical Practice and the Declaration of Helsinki principles for ethical research. The Institutional Review Board of the local academic institution and the children’s hospital from which samples were obtained approved the study protocol (IRB#2020-493). All authors reviewed the manuscript for accuracy, completeness of data and fidelity to the approved protocol.

### BLOOD SAMPLE COLLECTION AND PROCESSING

Blood samples were collected from the regional children’s hospital in Orleans Parish, Louisiana between March 18^th^-May 15^th^, 2020. A Stay At Home Order was issued on March 23rd and elective surgeries were halted on March 19th. The first pediatric case and death in Louisiana was reported on March 22^nd^ after autopsy confirmed SARS-CoV-2 infection.^15^ During the study period, due to hospital guidelines and state recommendations, in person, routine pediatric visits were limited. Samples therefore do not reflect the general population and likely include a higher proportion of children with chronic and/or life threatening conditions, time-sensitive surgical needs, acute medical, mental health, or trauma related concerns. Blood was drawn as part of care in the emergency room, inpatient floors, ambulatory clinics, or as part of routine pre-operative studies for time-sensitive surgeries. Due to hospital regulations, refrigerated samples were release to our study team seven days after blood draw. Serum or plasma was then immediately isolated, heat inactivated for 30 minutes at 56°C, and stored at −20°C until analyzed. All identifying data was removed and samples were coded with a unique subject identification.

### SARS-CoV-2 IgG ELISA

A serum immunoglobulin G (IgG) ELISA was used to determine presence or absence of SARS-CoV-2 anti-Spike (S) antibodies. Briefly, detergent-solubilized viral glycoproteins produced by infected or transfected cells were immobilized in wells coated with Concanavalin-A (ConA) to provide solid phase antigen for ELISA as previously described.^16^ Antigens were generated by transfection of 293T (ATCC) cells with SARS-CoV-2 S-protein mammalian expression plasmid (BEI Resources). ELISA plates were coated with 10 µg/ml ConA (Vector Labs). Antigen positive and negative cells were lysed and centrifuged to remove insoluble debris. Predetermined optimal dilutions of cell lysates were incubated in ConA coated wells to allow capture of viral glycoproteins, followed by incubation of serum samples diluted 1:100. Antibody binding was detected with peroxidase conjugated anti-human IgG (Jackson Immunoresearch) and the colorimetric reaction was performed as previously described.^16^ Each serum sample was tested in parallel with similarly prepared lysates of untransfected 293T cells with the latter optical density (OD) reading subtracted to determine net OD. Positive reactions were defined as a net OD reading >0.7. All antibody analyses were performed blind to all medical data including demographic data. This ELISA was validated by testing of COVID-19 convalescent serum and 100 pre-COVID-19 sera samples from healthy subjects collected prior to 2020.

### DATA SOURCES AND STATISTICS

The following data was abstracted from electronic medical records: demographics, chief complaint at the visit when the blood was drawn, diagnostic codes for current visit and chronic medical conditions, and history of COVID-19 testing. Median household income for the patient’s zip code of residence was used as a proxy for socioeconomic status (SES) and obtained from the US Census Bureau (2014-2018 averaged values) or from a commercial vendor that provides mined US Census Bureau data from 2018 (https://www.cubitplanning.com/). Parishes (counties) with >30 subjects were identified. Parish level COVID-19 case rates were extracted from Louisiana Department of Health website: ldh.la/gov/Coronavirus over the same period.

Statistical analysis was performed and graphs created using GraphPad Software v7 and IBM SPSS Statistics (v.25). Categorical measures are presented as percentages and continuous measures as medians and interquartile ranges. Analysis between seronegative and seropositive groups was performed using Chi squared tests. Map was created using Datawrapper (https://www.datawrapper.de/).

## Results

### DEMOGRAPHIC AND CLINICAL CHARACTERISTICS

A total of 1690 stored blood samples from pediatric subjects were obtained in New Orleans from March 18^th^ to May 15th, 2020 (Figure 1). From March 18^th^ through April 20^th^, only gold-top serum-separator tubes were collected. From April 21^st^ green (heparin) and lavender (EDTA) were collected, accounting for a higher volume of samples. Duplicate blood samples (from the same patient, on the same day) accounted for 130 of the 1690 samples. Additionally, 744 samples were from subjects that had blood collected on more than one day. In total, 812 unique subjects were tested during the study period by ELISA for serum IgG antibodies binding to SARS-CoV-2 Spike (S) protein. As shown in Figure 2, our study included subjects from 41 of 64 parishes within Louisiana. A majority of patients were from Jefferson (27.5%) or Orleans (27.1%) Parishes and fewer subjects were from St. Tammany, Tangipahoa, Terrebonne and St. Bernard Parishes.

**Figure 1.**
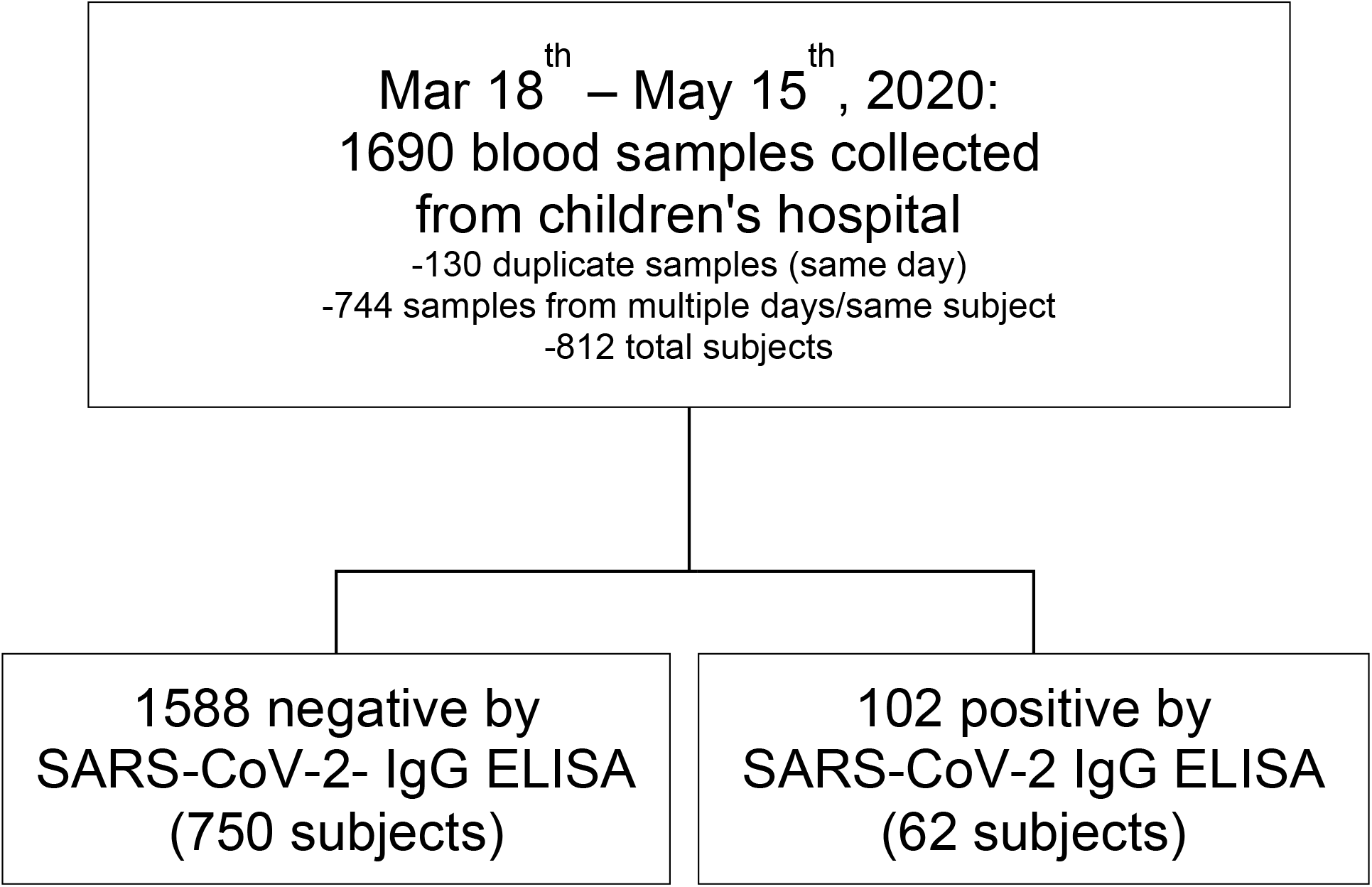
Overview of seroprevalence testing of pediatric blood samples.

**Figure 2.**
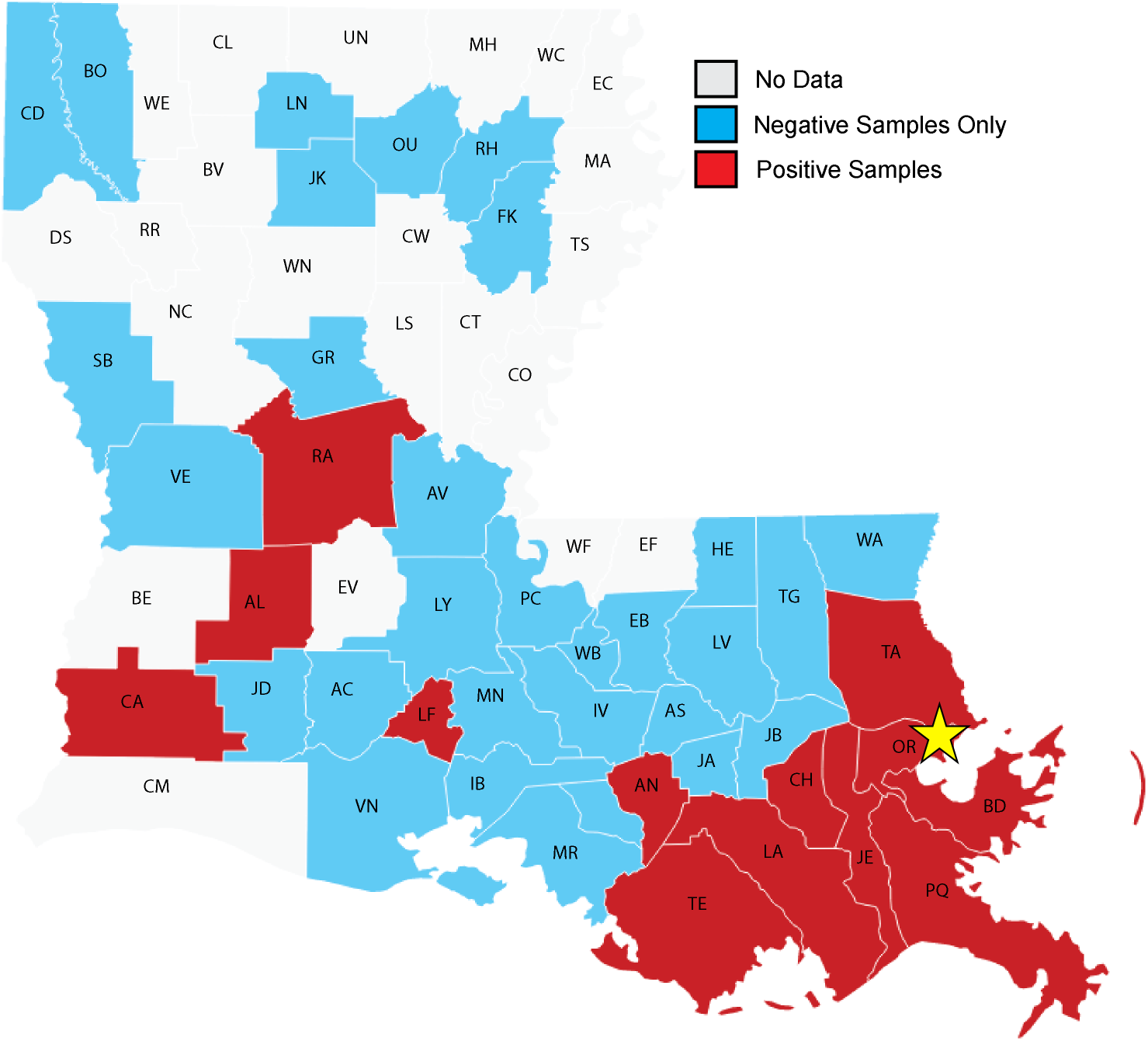
Map of Louisiana parishes represented in study subject blood samples collected from a children’s hospital during Stay At Home Order. Parishes are represented by individual colors with seropositive (red), seronegative (blue), or not represented (white) indicated. Yellow star indicates parish of hospital. (N=60, with patients from outside Louisiana not shown). Parishes with >30 subjects include OR (Orleans), JE (Jefferson), TE (Terrebonne), TA (St. Tammany), BD (St. Bernard), and TG (Tangipahoa). Complete list of abbreviations in Table S2.

The demographic characteristics of the patients are shown in Table 1. The population was evenly split between females (50.4%) and males (49.6%). The age range was 2 days to 18 years of age. The median age was 11 years (interquartile range (IQR), 4-15 years). Our cohort included 42.7% children who identified as Black, 41.4% as white, 5.3% as Hispanic, and 10.6% who identified as other races or race/ethnicity was not listed.

**Table 1.** Demographic Characteristics of Pediatric Populations During Louisiana Stay At Home Order.

| Characteristics | SARS-CoV-2 Serostatus | | | Significant $\chi^2$ |
| --- | --- | --- | --- | --- |
|  | Negative<br>(N=750) | Positive<br>(N = 62) | Total<br>(N = 812) |  |
| **Female sex - no. (%)** | 374 (49.9) | 35 (56.5) | 409 (50.4) | ns |
| **Age range** | 2 days - 18 yr | 2 mo - 18 yr | 2 days - 18 yr |  |
| **Age median (IQR)** | 11 yr (4-15) | 10 yr (4.75-15) | 11 yr (4-15) |  |
| 0-11 mo | 58 (7.7) | 3 (4.8) | 61 (7.5) | ns |
| 12-35 mo | 76 (10.1) | 7 (11.3) | 83 (10.2) |  |
| 36 mo - 5 yr | 92 (12.3) | 7 (11.3) | 99 (12.2) |  |
| 6-9 yr | 101 (13.5) | 13 (21.0) | 114 (14.0) |  |
| 10-14 yr | 226 (30.1) | 14 (22.6) | 240 (29.6) |  |
| 15-18 yr | 197 (26.3) | 18 (29.0) | 215 (26.5) |  |
| **Race - no. (%)** |  |  |  |  |
| Black | 311 (41.5) | 36 (58.1) | 347 (42.7) | P = 0.003 |
| White | 321 (42.8) | 15 (24.2) | 336 (41.4) |  |
| Hispanic | 36 (4.8) | 7 (11.3) | 43 (5.3) |  |
| Other | 82 (10.9) | 4 (6.5) | 86 (10.6) |  |
| **Parish - no. (%)** |  |  |  |  |
| Orleans | 191 (25.5) | 29 (46.8) | 220 (27.1) | P = 0.023 |
| Jefferson | 208 (27.7) | 15 (24.2) | 223 (27.5) |  |
| St Tammany | 50 (6.7) | 3 (4.8) | 53 (6.5) |  |
| St. Bernard | 29 (3.9) | 1 (1.6) | 30 (3.7) |  |
| Terrebonne | 32 (4.3) | 1 (1.6) | 33 (4.1) |  |
| Tangipahoa | 40 (5.3) | 0 (0.0) | 40 (4.9) |  |
| Other | 195 (26) | 13 (21) | 208 (25.6) |  |
| **Median household income\*** | \$49,895 | \$43,632 | \$49,838 | |
| **Median household income** |  |  |  |  |
| **Quartiles - no. (%)** |  |  |  |  |
| 0-25% (<\$36,939) | 179 (24.1) | 27 (43.5) | 206 (25.6) | P = 0.006 |
| 26-50% (\$36,939 - <49,838) | 190 (25.6) | 9 (14.5) | 199 (24.7) | |
| 50-75% (\$49,838 - <61,252) | 192 (25.8) | 15 (24.2) | 207 (25.7) | |
| 75-100% (\$61,252 - 107,546) | 182 (24.5) | 11 (17.7) | 193 (24.0) | |

\*residence area median household income estimated from zip code US Census data 2018.

Based on diagnostic codes from past medical history, 17.7% of seropositive subjects were categorized as having a severely immunocompromising condition, including malignancy, organ failure, and bone marrow or stem cell transplant (Table 2). The majority of subjects presented for a problem or follow-up directly related to an existing chronic condition (43.5%), while 9.7% presented for trauma, 9.7% for behavioral health reasons, and 27% with a complaint or reason not directly related to an existing chronic condition, trauma, or behavioral health (Table 2).

**Table 2.** Presenting Complaint and Pre-Existing Conditions at Blood Draw in Seropositive Subjects.

**SARS-CoV-2 seropositive patients**
| **Presenting Complaint - No (%)** | **All** | **Severe immunocompromising condition\*** |
| --- | --- | --- |
| **Complication of or follow-up for existing chronic condition** | 27 (43.5) | 9 (14.5) |
| **Complaint not directly related to an existing chronic condition, trauma, or behavioral health** | 17 (27.4) | 0 (0) |
| **Trauma** | 6 (9.7) | 0 (0) |
| **Behavioral Health** | 6 (9.7) | 0 (0) |
| **Other or unclear** | 6 (9.7) | 2 (3.2) |
| **Total** | 62 (100) | 11 (17.7) |

\*Severely immunocompromising existing conditions in this category include: any malignancy, end-stage renal disease, chronic liver failure, chronic respiratory failure, heart failure, preparation for or receipt of stem cell transplant or bone marrow transplant, requiring hemodialysis, total parenteral nutrition dependence, aplastic anemia, hemophagocytic lymphohistiocytosis, cystic fibrosis, personal history of extra-corporeal membranous oxygenation

Overall, 7.6% of the patients tested positive for IgG antibody to SARS-CoV-2 S protein (Table 1). The earliest positive sample was from March 19^th^, from a 10 year-old child. We observed an increase in SARS-CoV-2 seroprevalence over time from March 18th to May 15th (Fig 3A). There was no statistically significant difference in seroprevalence by sex or age. There were statistically significant differences by race, parish, and residence in an area with lower household incomes (estimated by zip code). Although Black and white children were equally represented in the full cohort, Black children accounted for 58.1% of seropositive subjects, while white children represented only 24.2% of the seropositive subjects. A similar disproportionate rate of seropositivity was also seen Hispanic children who comprised 5.3% of all subjects, but 11.3% of seropositive subjects. Positivity rates within race also differed with the lowest rate found in white children (4.5%), and significantly higher rates in Black children (10.4%) and Hispanic children (16.2%).

**Figure 3.**
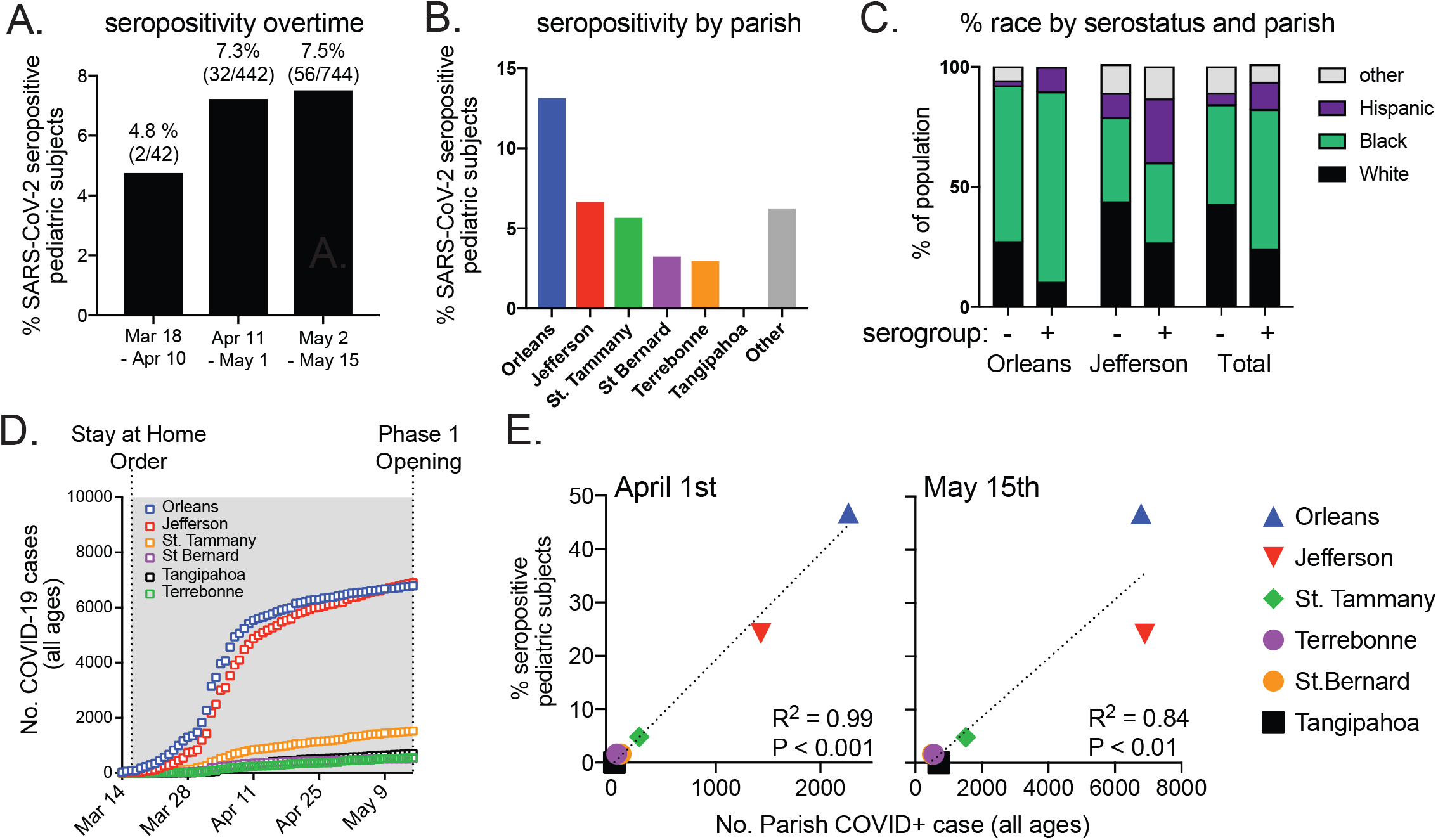
Seropositivity in pediatric subjects increases overtime and correlates to reported parish COVID-19 Cases. A) Seropositivity in blood samples from pediatric subjects by 3-week intervals. B) Seropositivity by parish in pediatric subjects (from Louisiana Parishes with 30+ subjects). C) % racial group within Orleans and Jefferson Parish populations by serostatus. D) Number of cumulative COVID-19 cases by Parish reported to the state during Louisiana Stay At Home Order. E) % of all seropositive pediatric subjects represented by each parish and number of reported cumulative COVID-19 cases by parish during Louisiana Stay At Home Order for either April 1^st^ or May 15^th^. Dotted line at linear regression with R squared and P-value indicated.

As with race, seroprevalence rates differed by parish (Fig 3C). Despite approximate overall equal numbers in children from Orleans and Jefferson Parish in the cohort, children living in Orleans Parish accounted for 46.8% of seropositive children, children from Jefferson Parish accounted for 24.2% and children living in other parishes accounted for 4.8%, 1.6% and 1.6% in St. Tammany, St. Bernard and Terrebonne Parishes respectively. Positivity rates within each parish also differed significantly, with 13% of participants in Orleans Parish seropositive, while only 6.7% of children in Jefferson Parish were positive and 5.7% in St. Tammany (Fig 3B-3C). From data reported by the State, Orleans and Jefferson Parish also had higher levels of cumulative COVID-19 case rates for all ages (Fig. 3D). Seropositivity in children broken down by parish was significantly and linearly correlated to number of state-reported COVID-19 cases on two different dates (Fig 3E). Of seropositive subjects, 43.5% lived in a zip code with a median household income in the bottom quartile (<$36,939) while the distribution of subjects in the seronegative sample was equal across income levels.

Multiple samples obtained on different days were available in a subset of participants. In three seronegative subjects, samples were available across a period of 4-6 weeks without evidence of seroconversion (Fig S1A). Four subjects seroconverted over a one-month period after initially testing negative (Figure S1B). Six subjects showed continued presence of antibody against SARS-CoV-2 over a one-month period (Figure S1C). Of the 62 seropositive subjects, 33 had a medical record of a COVID-19 test for viral presence or serum IgG (Table S1). Of the 33 tested, 6 had positive COVID-19 results. Two subjects initially tested positive for antibodies against SARS-CoV-2, then tested negative at later time points (Figure S2).

## Discussion

Our study utilized serologic methods to evaluate evidence of previous SARS-CoV-2 infection in children receiving medical care in New Orleans during the early part of the COVID-19 outbreak in Louisiana. We demonstrated 7.6% positive seroprevalence for SARS-CoV-2 in this population, similar to a recent study in adults.^17^ Residents in Orleans Parish, while only about one fourth of the total study population, accounted for almost half (46.8%) of the positive cases. This result is consistent with reported infection rates where Orleans Parish was an early epicenter of COVID-19 having one of the highest per capita rates of infections in the country in March. Notably our earliest detected seropositive individual was from March 19^th^, days prior to the March 24^th^ Louisiana Stay At Home Order and concurrent with the first reported COVID-19 positive infection case in a pediatric patient, indicating that pediatric infections were present prior to the first documented case. Importantly, we also found a trend of initial increasing seroprevalence, which plateaued in late April, likely due to the state mandated Stay At Home Order, changes in social distancing guidance and remote learning or closure of schools across the state.

Similar to a recent study in New Orleans that determined seroprevalence in adults^17^, our data suggest SARS-CoV-2 infection in children is common. Limited seroprevalence studies in adults in the US^18^ and globally^19-21^ are highly variable in characteristics and outcomes, with seroprevalence rates ranging from 1.5% (Santa Clara County) to 14.9% in New York State. Given wide differences in demographics and disease course across different geographic locations, it is difficult to compare our results with those of other studies. A recently released report from Seattle Children’s Hospital with a sample source similar to our study showed a seroprevalence of 1% in their population.^22^ More research is needed to better understand the differences in infection dynamics between adults and children and differences across geographic areas.

We observed that seropositivity rates in children mapped well to regions with high state-reported COVID-19 case rates (Fig 3E). Given the expected lag of seroconversion following acute infection, it is perhaps not surprising that the correlation was stronger to the number of state-reported COVID-19 cases (all ages) from April 1^st^ (R^2^ = 0.99) rather than May 15^th^ (R^2^ = 0.84), the end date of blood samples tested in this study. Interestingly, between April 1^st^ and May 15^th^ there were notable changes in state-reported parish COVID-19 case rates (e.g., increasing cases in Jefferson Parish and Tangipahoa); future studies in community wide seroprevalence may demonstrate these changes, albeit with a time lag.

Our results also implicate neighborhood income level as an additional factor contributing to risk of infection. Existing literature links zip code to myriad health outcomes, including mortality.^23^ We report here that children residing in zip codes with lower household income account for 43.5% of positive cases, yet only 25.4% of the total sample. This result has been identified as a key parameter in seroprevalence rates of other common viruses such as cytomegalovirus^24^ and is also consistent with preliminary data suggesting a higher transmission of SARS-CoV-2 in lower income neighborhoods that may be confounded with the reported high rates of infection in minority populations.^25^

In addition to potential differences due to SES, our results also indicate racial disparity in infections, with 10.4% of Black children and 16.2% of Hispanic children testing positive for antibody, compared to the 4.5% seropositivity rate in white children. This mirrors existing literature which have shown racial disparities in infection rates, including a recent study in New Orleans in adults.^26^

Despite the strengths of this study, there are limitations. The study population consists of children obtaining medical care during the beginning of the pandemic and is not likely to be representative of the general population. Our data on both race and SES were obtained from electronic medical records. In terms of SES, we utilized estimated average household income from existing data through 2018 and do not suggest that this is a true estimate of the individual SES of the family in which the child lives, but rather is an indicator of the established links between neighborhood factors and health risk. Race was also obtained from electronic medical records and not directly obtained from the participants, as such mixed race categorization is lacking. Our antibody ELISA only captures children who have mounted an antibody response, missing children who have been infected but fail to produce antibodies. It has been reported that some individuals do not produce detectable antibodies after SARS-CoV-2 infection^27^ and there may be short-term antibody persistence in some individuals. Lastly, the geographical location of the hospital would be most accessible to those residing in Orleans Parish. Children with severe medical emergencies residing in other parishes may have sought care at closer hospitals resulting in less representation of these regions.

An enhanced understanding of the general seroprevalence in pediatric populations is critical to implementing public safety protocols related to schooling, daycare and other activities that involve children and families. Our data indicate that children are susceptible to SARS-CoV-2 infection and develop measurable antibodies, and that existing racial and socioeconomic disparities seen in adults are reflected in children. Our overall seroprevalence of 7.6% clearly shows that mitigation measures at schools and interventions targeting children’s social interactions may be crucial to control viral spread.

## Data Availability

All data can be obtained through corresponding author upon publication

## Author contributions

MLD, EBN, SSD, JSS, JER, KJZ designed the study and have full access to all data in this study. MLD, EBN, DE, AS, JR, NGB, KAM, AP, RK, AS, TSH, SS, AES, RC, AL, LS, RU, SSD, JER, KJZ were responsible for sample and medical data collection, sample processing, and data processing. MLD, EBN, SSD, JSS, JER, KJZ contributed to writing the manuscript. MLD, EBN, LM, SSD, JER, KJZ had roles in data collection, data analysis and data interpretation. MLD, EBN, LM completed all statistical analysis. All authors contributed to final manuscript review and approval.

## Acknowledgements

We thank our department chairs Drs. Chad Steele and Samir El-Dahr for their encouragement and financial support on this project; Dr. Joshua Yukich for insightful comments; Children’s Hospital New Orleans and the CHNOLA Clinical Trials Center; the CHNOLA clinical laboratory staff, and the numerous doctors, nurses and families who are helping overcome this pandemic.

